# “Similar performances but markedly different triaging thresholds in three CAD4TB versions risk systematic errors in TB screening programs”

**DOI:** 10.1101/2022.04.29.22274472

**Authors:** Jana Fehr, Emily B. Wong

**Affiliations:** Digital Health & Machine Learning, Hasso Plattner Institute for Digital Engineering and University of Potsdam, Germany; Africa Health Research Institute, KwaZulu-Natal, South Africa; Harvard Medical School, Boston, MA, USA; Division of Infectious Diseases, Massachusetts General Hospital, Boston, MA, USA; Division of Infectious Diseases, University of Alabama at Birmingham, AL, USA

## Abstract

Tuberculosis (TB) screening programs may apply computer-aided diagnostic (CAD) tools on chest radiographs to select people for microbiological sputum examination using a pre-selected triaging threshold. CAD software-updates are frequently introduced and it is unknown whether their use requires adjustment of triaging thresholds. In a community-based screening program in South Africa, we compared the scores between the three recent CAD4TB versions (v5, v6, and v7) and assessed their performance to identify microbiologically-confirmed TB. The performance of all versions was similar (v5: AUC 0.78, v6: AUC 0.79, v7: AUC 0.80; p-values>0.05), but along a 0-100 point scale, each had markedly different score distributions and optimal triaging thresholds (v5: 40, v6: 47, v7: 20). This has the potential to cause confusion within TB screening programs as these tools are increasingly adopted and new versions released. Independent guidance for adapting CAD triaging thresholds for frequently released software updates is needed.

---

In a 2021 update, the World Health Organization (WHO) recommended TB screening programs in high burden communities to use chest radiography paired with computer-aided detection (CAD) tools to selectively target individuals who require microbiological sputum assessment for *Mycobacterium tuberculosis* (*Mtb*).^1^ The CAD-software CAD4TB (©Delft) quantifies increasing lung field abnormalities suggestive of active TB with a score between 0-100. Use of CAD4TB requires screening programs to select a triaging threshold above which participants are referred for microbiological sputum testing. CAD4TB is updated annually and the current version is the 7^th^ to be released (v7). Screening programs might be eager to use new versions because they offer improved performance^2^ or new utility features. However, it is unclear which steps are needed to evaluate and adjust screening procedures to new software versions and whether each new version requires setting a new triage thresholds.

Previous studies showed that triaging thresholds are not universal and require careful adjustment to demographic characteristics, laboratory capacities, budget, healthcare settings and study goals.^2,3,12,13,4–11^ Multiple studies compared the performance among different CAD-software platforms,^11,14,15^ but comparisons between software versions are lacking in the literature and are needed to support screening programs when using CAD-software and negotiating software updates.

We previously evaluated the performance of CAD4TBv5 and v6 in a community-based screening program in rural South Africa^12^ and here sought to assess the performance of the most recently released version CAD4TBv7 in the same collection of chest x-rays. As previously described, between 2018 and 2019, we used CAD4TB version 5 to triage participants for sputum examination (Xpert Ultra and Mtb culture) using a threshold of CAD4TBv5 25. Among the 9,912 participants who underwent chest radiography, n=5,594 (56.4%) were referred for sputum testing in the camp. A total of 99 (1.0%) participants had microbiologically positive sputum (either Xpert Ultra or Mtb culture positive), of which 75 (0.76%) participants were in the more stringent group excluding participants with only XpertUltra “trace” evidence.

We found that the overall performance to detect microbiologically confirmed TB on CXRs was similar between CAD4TB versions 5, 6 and 7 (AUC v5: 0.78, 95%CI [0.73, 0.83]; v6: 0.79 95%CI [0.73-0.84]; v7: 0.80 95%CI [0.75, 0.85]; p-value>0.1; Figure 1a). In a sensitivity analyses excluding those with only Xpert Ultra trace evidence, CAD4TBv7 showed slightly better performance than CAD4TBv5 (p-value=0.02) and CAD4TBv6 (p-value=0.04) to detect participants with TB (AUC v5: 0.82 [0.77, 0.87], v6: 0.84 [0.79, 0.89], v7: 0.86 [0.82, 0.91], Supplementary Figure 1).

**Figure 1:**
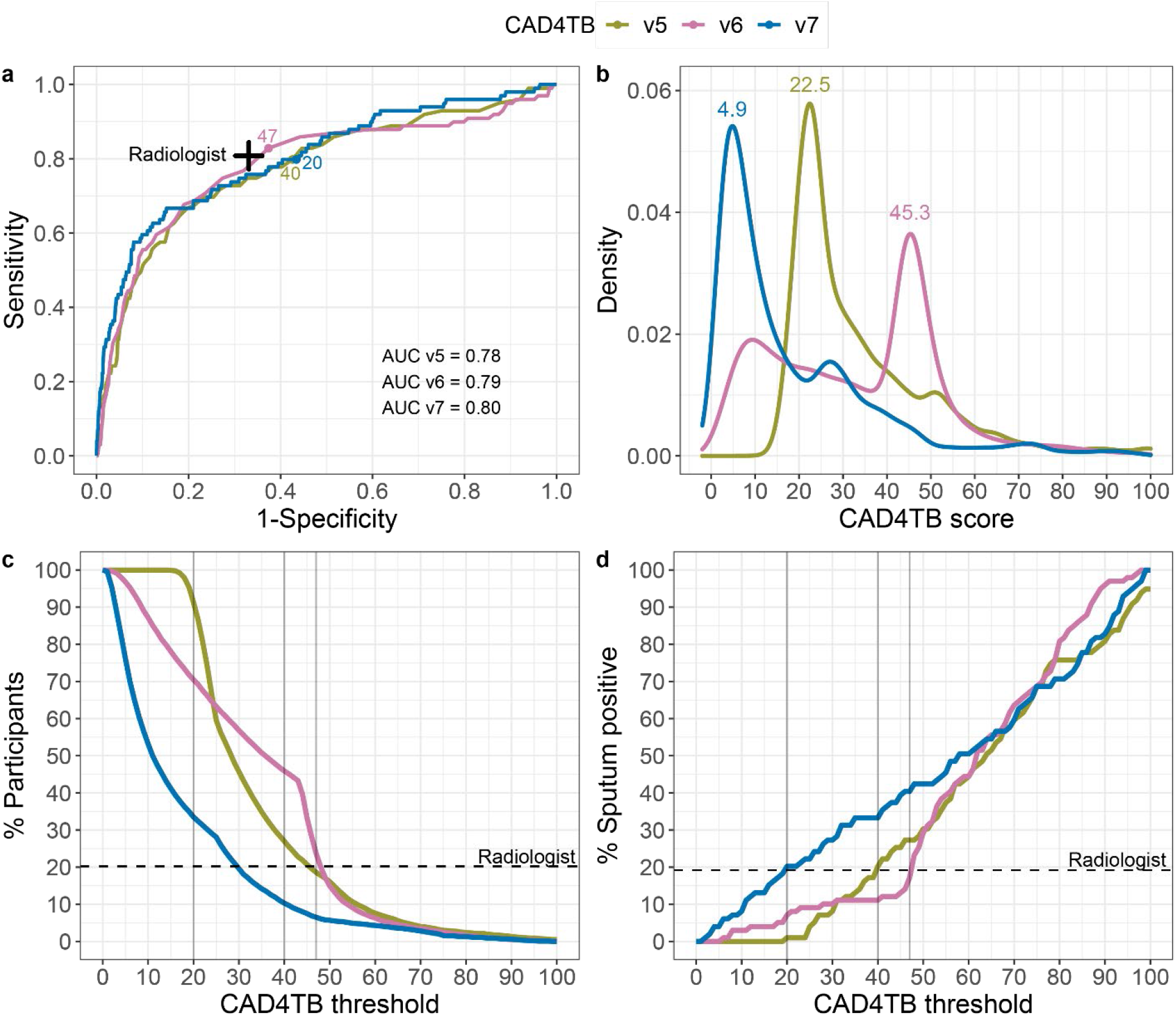
Performance of CAD4TB versions 5, 6 and 7 to identify microbiologically-confirmed tuberculosis (TB). TB was defined if sputum was found positive with either Xpert Ultra or microbiological culture. Figure a: Performance is depicted as sensitivity and specificity and area under the receiver-operating curve (AUC). Annotations show thresholds, which closest matched the radiologist’s sensitivity. Figure b shows distributions and most frequent CAD4TB scores of all three versions. Figures c and d show the percentage of participants triaged for sputum testing (c) and percentage of missed positive sputum (d) at each CAD4TB threshold. The performance of the senior radiologist is marked with a cross (a) and dashed lines (c, d).

We found that the distribution of CAD4TB scores varied significantly between the three most recent versions of the software with the scores of CAD4TBv7 significantly lower than in previous versions (Figure 1b): Median score for v5 (28, IQR 22-41), v6 (35, IQR 16-46), and v7 (11, IQR 5.2-27), p-value<0.001; Figure 1). No threshold from any version was able to match the performance target by the WHO of ≥90% sensitivity and ≥70% specificity.^16^ Therefore, we identified one threshold for each CAD4TB version that most closely matched the radiologist sensitivity at 80.8% [95%CI 71.7-88.0%] (Supplementary Table 1). The matching thresholds were v5: 40 (79.8% [95%CI 70.5-87.2%]), v6: 47 (82.8% [95%CI 73.9-89.7]), and v7: 20 (79.8 [95%CI 70.5-87.2]). At these thresholds, the three CAD4TB versions had slightly lower specificity than the radiologist (radiologist: 66.9% [95%CI 65.6-68.2]; v5 40: 57.4 [95%CI 56.0-58.8]; v6 47: 62.6 [95%CI 61.2-64.0]; v7 20: 56.6 [95%CI 55.2-58.0]).

The percentage of participants who would qualify for sputum microbiological testing varied substantially among CAD4TB versions for all potential triaging scores below 50 (Figure 1c, Supplementary Table 2). The same substantial variation was observed regarding the number of cases of microbiologically positive sputum that would be ‘missed’ using potential triaging thresholds for the different CAD4TB versions (Figure 1d). For example, triaging with CAD4TB threshold 40, would result in sputum testing for 27.0% (v5), 45.9% (v6), and 10.3% (v7) of participants. At the same threshold, the percentage of missed microbiologically-confirmed TB cases would be 20.2% (v5), 11.1% (v6), and 33.3% (v7).

The overall performance to detect microbiologically-confirmed TB was similar between CAD4TB versions 5, 6, and 7, but the distribution of scores across the 100 point scale varied greatly across the three versions. This meant that each numerical threshold had strikingly different performances across versions. To deal with such intra-version variation, screening programs will need to select new triaging thresholds for each new software update. Previous work^9,12^ and the developer^17^ suggest conducting a population-specific pilot study to finding triage thresholds that optimally serve the goals of each screening exercise. The design and conduct of such pilots, however, may result in additional costs, slow timelines and require alterations to established screening workflows.

It is unclear if each software update requires a new piloting exercise for re-adjustment or if re-calibration of each program’s triage threshold can be achieved through retrospective analysis of the newest version’s performance against population-specific CXR collections. For the community of end-users (e.g. those designing TB screening programs) decisions about programmatic adjustments to new versions are especially difficult because the underlying algorithmic or data changes between software versions are not communicated by manufacturers. Changes to the underlying TB reference standard for training might impact CAD-scores and require re-adjustment of triaging thresholds, whereas small changes for faster radiograph interpretation, might not. These important information, however, are not transparently shared with the community as CAD-developing companies have considered details about the geographic and clinical features of training data to be proprietary.^17^

Our performance estimates of CAD4TB are lower compared to other studies^2,14^. One possible explanation for this is that other studies have enrolled symptomatic patients seeking TB diagnosis, whereas our study was a community-screening, which enrolled participants regardless of symptom status, which revealed more subclinical and early-stage TB.^16^ Limitations of our study have been described^12^ and include that only people with symptoms or a CAD4TBv5 score greater or equals 25 were selected for microbiological sputum assessment, which might have biased our performance assessment results. A limitation of this study, is that our analysis is limited to a single CAD software. It is unknown whether our findings of significant variation between CAD4TB versions is applicable to other image interpretation algorithms used for TB screening.^17^

The wide variation of triaging thresholds between different software versions poses a hazardous risk of end-users introducing systematic errors. Communicating changes between software-versions and clear guidance for medical or public health users is urgently needed. End-users require transparency and guidance to successfully negotiate the rapidly changing landscape of software outputs and to effectively use CAD-tools for TB screening programs.

## Methods

### Study design

During a multi-disease community-based screening program in a rural district in KwaZulu-Natal, South Africa, local residents from the age of 15 were invited to participate in a free health check-up at a mobile camp (Details are described elsewhere^12,19^). From 25 May 2018 until May 2019 9,912 participants answered questions about current symptoms and underwent digital posterior-anterior chest x-ray (CXR) imaging using a mobile scanner unit (Canon CXDI-NE). Following WHO guidelines for TB prevalence surveys^20^, participants were referred for sputum examination if they reported any current TB-related symptom (fever, weight loss, cough, or night sweats) or if their CXR showed lung field abnormality. CXRs were analysed in real-time on a local workstation using CAD4TB version 5 (CAD4TBv5) and scored between 0-100 to indicate the likelihood of TB-related lung field abnormality. An initial pilot phase determined CAD4TBv5 score at 25 as optimal threshold to classify abnormal CXRs in this screening setting and those with CAD4TBv5 equal or higher 25 were referred for sputum testing. CAD4TB version 6 (CAD4TBv6) and version 7 (CAD4TBv7) scores were retrospectively calculated. A senior radiologist interpreted CXRs blinded to CAD4TB scores and any patient information as having either normal or abnormal lung fields. Sputum specimens were analysed for *Mycobacterium tuberculosis* (Mtb) using Xpert Ultra MTB/RIF® (XpertUltra) and liquid BACTEC MGIT culture (MGIT).

### Data analysis

We analysed data from 9,912 participants who completed chest radiography. Sputum was defined positive if either XpertUltra or MGIT were positive for *Mtb*. We performed a sensitivity analysis with an additional, more stringent diagnostic group, which excluded participants with “trace” XpertUltra results and negative MGIT.

We compared the median, interquartile range (IQR) and distribution peaks between scores calculated with CAD4TB versions 5, 6 and 7 using paired Wilcoxon-rank tests. We assessed the performance of CAD4TB scores to identify microbiologically-confirmed TB on CXRs by generating receiver operating characteristic (ROC) curves and then comparing the areas under the curve (AUC) using the DeLong test.^21^ For each threshold, we calculated 1) sensitivity, specificity, negative predictive value (NPV), positive predictive value (PPV) with 95% confidence intervals (CI), 2) the percentage of participants above the triaging thresholds that would prompt sputum testing 3) the percentage of participants with microbiologically positive sputum who were missed by the triage threshold and 4) the number needed to test (NNT) to identify one participant with *Mtb* positive sputum. All statistical analyses were performed using R (version 4.1.2).

## Data Availability

The Vukuzazi screening protocol as well as the dataset analyzed during the current study may be accessed via the AHRI Data Repository at https://data.ahri.org/index.php/catalog/990 upon approval of proposed analyses by the Vukuzazi Scientific Steering Committee and completion of a data access agreement.

## Author Contributions

JF performed data analysis with figures and tables and writing of the report. EBW contributed to study design, analysis conceptualization, revision and supervision. Both authors approved the final version.

## Notes

### Competing Interest Statement

The authors have declared no competing interest.

### Funding Statement

The community-screening program 'Vukuzazi' is funded by the Africa Health Research Institute, the Wellcome Trust (201433/Z/16/Z), and the Bill and Melinda Gates Foundation (OPP1175182). Additional funders include NIAID (K08AI118538) and FIC (TW011687), National Institutes of Health, and Cascade IMPAc-TB Center (Contract # 75N93019C00070). The funders had no role in study design, data analysis and interpretation, or writing of the report.

### Author Declarations

Ethics committees of the University of KwaZulu-Natal Biomedical Research Ethics Committee (BE560/17), the London School of Hygiene & Tropical Medicine Ethics Committee (14722), and the Partners Institutional Review Board (2018P001802) gave ethical approval for this work.

## References

1. World Health Organization. WHO consolidated guidelines on tuberculosis. Module 2: Screening Systematic screening for tuberculosis disease. Who (2021).

2. Murphy, K. et al. Computer aided detection of tuberculosis on chest radiographs: An evaluation of the CAD4TB v6 system. in arXiv 1–11 (2019) doi:10.1038/s41598-020-62148-y.

3. Muyoyeta, M. et al. The sensitivity and specificity of using a computer aided diagnosis program for automatically scoring chest X-rays of presumptive TB patients compared with Xpert MTB/RIF in Lusaka Zambia. in PLoS One 9, 16–18 (2014).

4. Breuninger, M. et al. Diagnostic accuracy of computer-aided detection of pulmonary tuberculosis in chest radiographs: A validation study from sub-Saharan Africa. in PLoS One 9, (2014).

5. Rahman, M. T. et al. An evaluation of automated chest radiography reading software for tuberculosis screening among public-and private-sector patients. in Eur. Respir. J. 49, (2017).

6. Koesoemadinata, R. C. et al. Computer-assisted chest radiography reading for tuberculosis screening in people living with diabetes mellitus. in Int. J. Tuberc. Lung Dis. 22, 1088–1094 (2018).

7. Philipsen, R., Ginneken, B. Van & Melendez, J. Computer Aided Detection of Tuberculosis CAD4TB. (2018).

8. Melendez, J. et al. Automatic versus human reading of chest X-rays in the Zambia National Tuberculosis Prevalence Survey. in Int. J. Tuberc. Lung Dis. 21, 880–886 (2017).

9. Qin, Z. Z. et al. Using artificial intelligence to read chest radiographs for tuberculosis detection: A multi-site evaluation of the diagnostic accuracy of three deep learning systems. in Sci. Rep. 1–10 (2019) doi:10.1038/s41598-019-51503-3.

10. Zaidi, S. M. A. et al. Evaluation of the diagnostic accuracy of Computer-Aided Detection of tuberculosis on Chest radiography among private sector patients in Pakistan. in Sci. Rep. 8, 1–9 (2018).

11. Khan, F. A. et al. Articles Chest x-ray analysis with deep learning-based software as a triage test for pulmonary tuberculosis : a prospective study of diagnostic accuracy for culture-confirmed disease. in Lancet Digit. Heal. 2, e573–e581 (2020).

12. Fehr, J. et al. Computer-aided interpretation of chest radiography reveals the spectrum of tuberculosis in rural South Africa. in npj Digit. Med. 4, 20 (2021).

13. Nishtar, T., Burki, S., Ahmad, F. S. & Ahmad, T. Diagnostic accuracy of computer aided reading of chest x-ray in screening for pulmonary tuberculosis in comparison with Gene-Xpert. in Pakistan J. Medicical Sci. 38, (2022).

14. Qin, Z. Z. et al. Tuberculosis detection from chest x-rays for triaging in a high tuberculosis-burden setting: an evaluation of five artificial intelligence algorithms. in Lancet Digit Heal. 3, 543–54 (2021).

15. Kik, S. V. et al. Diagnostic accuracy of chest X-ray interpretation for tuberculosis by three artificial intelligence-based software in a screening use-case: an individual patient meta-analysis of global data. in medRxiv 10, 2022.01.24.22269730 (2022).

16. World Health Organization. High-priority target product profiles for new tuberculosis diagnostics: report of a consensus meeting. 1–96 (2014).

17. Qin, Z. Z. et al. A new resource on artificial intelligence powered computer automated detection software products for tuberculosis programmes and implementers. in Tuberculosis 127, (2021).

18. Wong, E. B. It is time to focus on asymptomatic tuberculosis. Clin. Infect. Dis. 1–27 (2020).

19. Wong, E. B. et al. Convergence of infectious and non-communicable disease epidemics in rural South Africa: a cross-sectional, population-based multimorbidity study. in Lancet Glob. Heal. 9, e967–e976 (2021).

20. World Health Organisation. Systematic screening for active tuberculosis: principles and recommendations. (2013).

21. DeLong, E. R., DeLong, D. M. & Clarke-Pearson, D. L. Comparing the Areas under Two or More Correlated Receiver Operating Characteristic Curves: A Nonparametric Approach. in Biometrics 44, 837 (1988).

