## Supplementary Material for "“Similar performances but markedly different triaging thresholds in three CAD4TB versions risk systematic errors in TB screening programs”"

**Supplementary Figure 1:** Receiver operating curves of CAD4TB versions 5, 6 and 7 to detect microbiologically-positive sputum on chest radiographs. Positive was defined as positive if either Xpert Ultra or culture was positive, excluding participants who had a Xpert trace and negative culture results. Annotations show score thresholds, which closest matched the radiologist's sensitivity (cross).

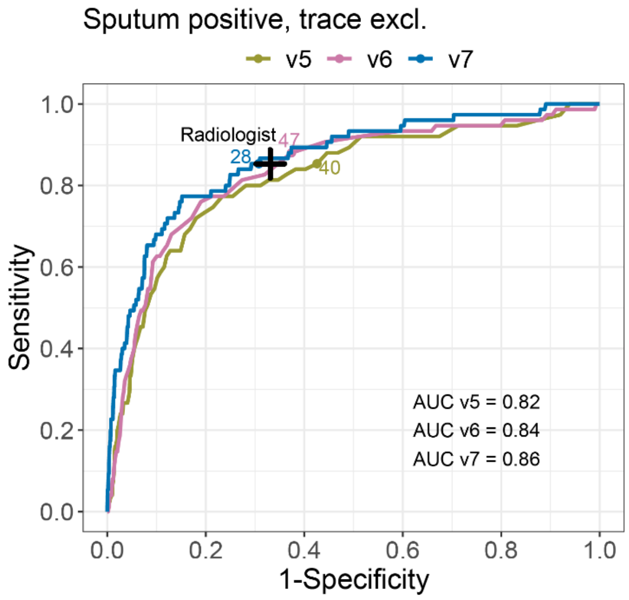

**Supplementary table 1:** Performance of radiologist to detect microbiologically-confirmed TB on chest radiographs and CAD4TB scores of versions 5, 6 and 7, which match the sensitivity of the radiologist. Performance is given with sensitivity, specificity, positive predictive value (PPV), negative predictive value (NPV), percentage of participants that is triaged for sputum testing, percentage of people with positive sputum that are missed, and number of participants that need to be tested to find one person with positive sputum (NNT). Sensitivity, specificity, PPV, and NPV estimates are given with 95% confidence intervals (CI).

| | **Sensitivity % (CI)** | **Specificity % (CI)** | **PPV % (CI)** | **NPV % (CI)** | **% triaged for sputum testing (absolute n)** | **% Missed positive sputum** | **Number needed to test (NNT)** |
|---|---|---|---|---|---|---|---|
| **Radiologist: Any lung field abnormality** | 80.8 (71.7-88.0) | 66.9 (65.6-68.2) | 4.7 (3.8-5.8) | 99.4 (99.1-99.7) | 20.2 (2002) | 19.2 | 25 |
| **CAD4TBv5: 40** | 79.8 (70.5-87.2) | 57.4 (56.0-58.8) | 3.7 (2.9-4.5) | 99.3 (98.9-99.6) | 27.0 (2677) | 20.2 | 34 |
| **CAD4TBv6: 47** | 82.8 (73.9-89.7) | 62.6 (61.2-64) | 4.3 (3.4-5.3) | 99.4 (99.1-99.7) | 23.7 (2348) | 17.2 | 29 |
| **CAD4TB v7: 20** | 79.8 (70.5-87.2) | 56.6 (55.2-58.0) | 3.6 (2.9- 4.5) | 99.3 (98.9-99.6) | 33.5 (3324) | 20.2 | 42 |

**Supplementary table 2:** Performance of CAD4TB versions 5, 6, and 7 at the thresholds 10, 20, 30, 40, 50, and 60 to detect microbiologically-confirmed TB on chest x-rays. Performance is given with sensitivity, specificity, percentage of participants that is triaged for sputum testing, percentage of people with positive sputum that are missed, and number of participants that need to be tested to find one person with positive sputum (NNT). Sensitivity, specificity estimates are given with 95% confidence intervals (CI).

| threshold | CAD4TB Version | Sensitivity (CI) | Specificity % (CI) | % triaged for sputum | % missed positive sputum | Number needed to test (NNT) | PPV | NPV |
|---|---|---|---|---|---|---|---|---|
| 10 | V5 | 100 (96.3-100) | 0 (0-0.1) | 100 | 0 | 100 | 2.0 (1.6-2.4) | Not estimable, because no scores <10 |
| | V6 | 97.0 (91.4-99.4) | 3.1 (2.6-3.6) | 87.2 | 3.0 | 90 | 2.0 (1.6-2.4) | 98.1 (94.4-99.6) |
| | V7 | 91.9 (84.7-96.4) | 38.4 (37.0-39.8) | 53.2 | 8.1 | 58 | 2.9 (2.4-3.6) | 99.6 (99.2-99.8) |
| 20 | V5 | 99.0 (94.5-100) | 1.4 (1.1-1.7) | 91.3 | 1.0 | 92 | 2.0 (1.6-3.4) | 98.5 (92.1-100) |
| | V6 | 92.9 (86.0-97.1) | 11.7 (10.8-12.6) | 70.5 | 7.1 | 76 | 2.1 (1.7-2.6) | 98.8 (97.5-99.5) |
| | V7 | 79.8 (70.5-87.2) | 56.6 (55.2-58.0) | 33.5 | 20.2 | 42 | 3.6 (2.9-4.5) | 99.3 (98.9-99.8) |
| 30 | V5 | 91.9 (84.7-96.4) | 28.7 (27.5-30.0) | 45.7 | 8.1 | 50 | 2.6 (2.1-3.1) | 99.4 (98.9-99.8) |
| | V6 | 89.9 (82.2-95.0) | 22.4 (21.2-23.6) | 56.8 | 10.1 | 63 | 2.3 (1.8-2.8) | 99.1 (98.3-99.6) |
| | V7 | 72.7 (62.9-81.2) | 72.8 (71.6-74.1) | 19.6 | 27.3 | 27 | 5.2 (4.1-6.4) | 99.2 (98.9-99.5) |
| 40 | V5 | 79.8 (70.5-87.2) | 57.4 (56.0-58.8) | 27.0 | 20.2 | 34 | 3.7 (2.9-4.5) | 99.3 (98.9-99.6) |
| | V6 | 88.9 (81.0-94.3) | 33.3 (31.9-34.6) | 45.9 | 11.1 | 52 | 2.6 (2.1-3.2) | 99.3 (98.8-99.7) |
| | V7 | 66.7 (56.5-75.8) | 84.6 (83.6-85.6) | 10.3 | 33.3 | 15 | 8.1 (10.2-99.2) | 99.2 (98.9-99.5) |
| 50 | V5 | 69.7 (59.6-78.5) | 74.5 (73.3-75.8) | 16.1 | 30.3 | 23 | 5.3 (4.1-6.6) | 99.2 (98.8-99.4) |
| | V6 | 70.7 (60.7-79.4) | 76.4 (75.1-77.5) | 14.9 | 29.3 | 21 | 5.7 (4.5-7.2) | 99.2 (98.9-99.5) |

| | | | | | | | | |
|---|---|---|---|---|---|---|---|---|
| | V7 | 57.6 (47.2-67.5) | 91.4 (90.6-92.2) | 5.7 | 42.4 | 10 | 11.9 (9.2-15.2) | 99.1 (98.7-99.3) |
| 60 | V5 | 55.6 (45.2-65.5) | 87.9 (87.0-88.8) | 7.6 | 44.4 | 14 | 8.6 (6.5-11.0) | 99.0 (98.6-99.3) |
| | V6 | 55.6 (45.2-65.5) | 89.9 (89.0-90.7) | 6.3 | 44.4 | 11 | 10.1 (7.7-12.9) | 99.0 (98.7-99.3) |
| | V7 | 49.5 (39.3-59.7) | 93.5 (92.8-94.2) | 4.3 | 50.5 | 9 | 13.4 (10.0-17.3) | 98.9 (98.6-99.2) |
| 70 | V5 | 40.4 (30.7-50.7) | 93.5 (92.8-94.2) | 4.0 | 59.6 | 10 | 11.3 (8.2-15.0) | 98.7 (98.4-99.0) |
| | V6 | 36.4 (26.9-46.6) | 94.6 (93.9-95.2) | 3.5 | 63.6 | 10 | 12 (8.5-16.2) | 98.7 (98.3-99.0) |
| | V7 | 40.4 (30.7-50.7) | 95.8 (95.2-96.3) | 2.8 | 59.6 | 7 | 16.3 (11.9-21.6) | 98.8 (98.4-99.0) |
